# Modeling the Covid-19 Epidemic Using Time Series Econometrics

**DOI:** 10.1101/2020.06.01.20118612

**Authors:** Adam Goliński, Peter Spencer

**Author notes:** Department of Economics and Related Studies, University of York, YO10 5DD; (Adam Goliński). Department of Economics and Related Studies, University of York, YO10 5DD; (Peter Spencer).

## Abstract

The classic ‘logistic’ model has provided a realistic model of the behavior of Covid-19 in China and many East Asian countries. Once these countries passed the peak, the daily case count fell back, mirroring its initial climb in a symmetric way, just as the classic model predicts. However, in Italy and Spain, and now the UK and many other Western countries, the experience has been very different. The daily count has fallen back gradually from the peak but remained stubbornly high. The reason for the divergence from the classical model remain unclear. We take an empirical stance on this issue and develop a model that is based upon the statistical characteristics of the time series. With the possible exception of China, the workhorse logistic model is decisively rejected against more flexible alternatives.

## 1 Introduction

There are many different ways of analyzing and projecting the progress of an epidemic like Covid-19. The government and its advisors mainly rely upon large computer models that analyze the spread of the disease and the effects of public health interventions in fine detail (Ferguson et al. (2020)). These so-called ‘mechanistic’ models are largely theory-based and in that sense resemble the large theory-based models constructed for example by the Bank of England to analyze the effect of policy interventions on the economy. Other epidemiologists fit curves to the time series data and use the theoretical dynamics to make data-based predictions (Batista (2020), Jia, Li, Jiang, Guo, and Zhao (2020), Murray (2020)). Avery, Bossert, Clark, Ellison, and Ellison (2020) classify these models as ‘phenomenological’ and note their resemblance to reduced-form econometric models.

These approaches complement each other nicely. Large-scale theoretical models are very useful for analyzing policy interventions and other structural changes but can miss important links, especially when confronted with ‘black swan’ events such as a financial breakdown or the outbreak of an unknown virus like Covid-19. Small data-based models can provide better forecasts. The Monetary Policy Committee uses both types of model for informing their decisions and forecasting (Burgess et al. (2013)). Reduced form models can also be used to check the properties of the theoretical models and match them better to the data (Meenagh, Minford, and Wickens (2009)).

Although epidemiological models may differ in many respects, they are all based on the same theory and they invariably predict that the daily counts for infections and deaths follow a bell-shaped curve. In other words, once the peak is passed and the daily count begins to fall, it follows a path that mirrors the upward climb, before slowing to a halt. In the large models, this classic pattern follows from the dynamics of the epidemic, which naturally slows as the disease runs through the population and immunity increases. Behavioral feedbacks and policy interventions may also be important in slowing the spread of the disease. In the time series models, the daily count follows a bell-shaped path by assumption and means that the cumulative count follows an *S*—shaped logistic curve. This idea is at the center of policymaking in this area and expressions like ‘flattening the curve’ are now part of everyday conversation.

These symmetric dynamics have provided a reliable way of modelling outbreaks of influenza and other epidemics in the past. Indeed, simple regression models based on fitting the bell (or logistic) curve to the data, also proved accurate in predicting the path of the Covid-19 outbreak in China and many East Asian countries (Jia, Li, Jiang, Guo, and Zhao (2020), Batista (2020)). However, the experience of Italy and Spain, which is now being followed by the UK and many other countries, has been very different. The daily mortality figures have fallen back gradually from the peak in these countries, but have remained stubbornly high. This contrast is stark in the daily series plotted for China and for Italy and Spain in Figure 1.

**Figure 1:**
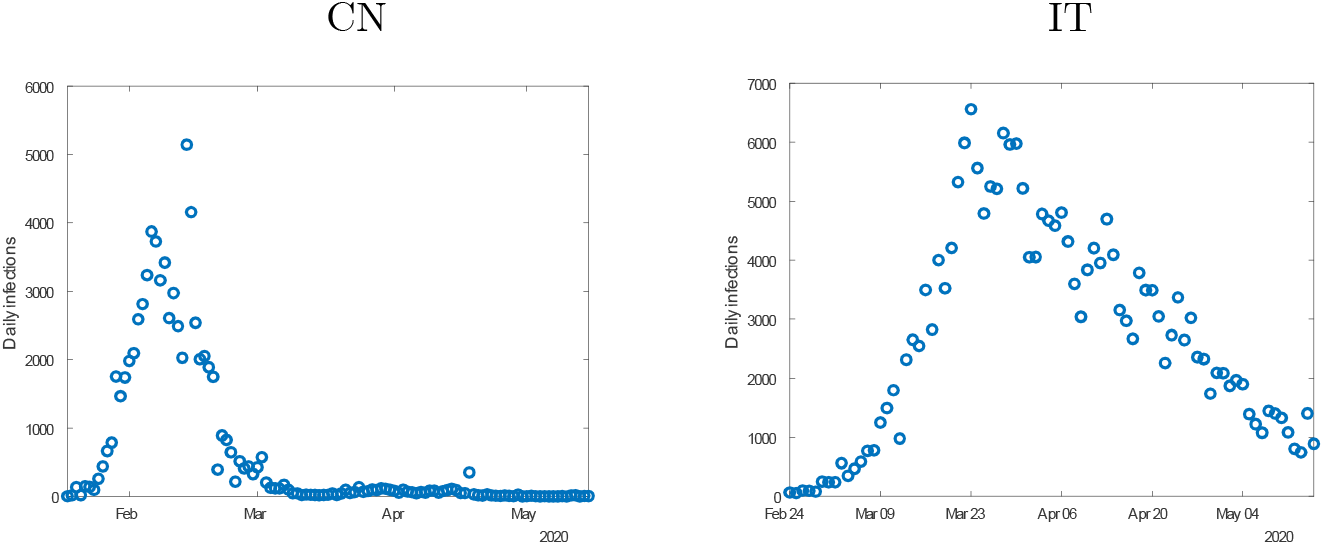
Covid-19 infections in China and Italy.

A positive skew in the national time series can appear because they aggregate data for areas that are hit by the virus at different times. However, US data for hospital admissions and fatalities areas like New York and New Jersey that have been badly affected by the virus exhibit a pronounced skew.^1^ Mortality data for English regions, which are arranged by date of death rather than date of announcement, thus avoiding the effect of skewed reporting delays, also exhibit a pronounced upper tail.^2^ Data for large US cities and English regions could also be prone to aggregation effects, but this seems less likely than for national aggregates.

One reason for the mortality figures to exhibit an upper tail is because the length of time from infection to death or recovery follows a gamma distribution and is positively skewed, as is well known (Hogg (1978), Bird (2013)). Some people recover very quickly, but others take much longer. This distribution is used by the *MRC* Biostatistics Unit to infer the true number of infections and the reproduction number (*R*) from mortality figures for the UK regions (Seaman and De Angelis (2020)). The regional breakdown helps deal with the problem that the epidemic affects regions at different times. This methodology also takes into account the positive skew in the reporting lag (Bird and Neilsen (2020), Birrell, Blake, van Leeuwen, and De Angelis (2020)).

There is also a positive skew in the infections data for Italy and many other countries. This is much harder to rationalize. Figure 1 contrasts the pattern of infections seen in China with that in Italy. We also find a positive skew in data for infections in Lombardy, the Italian region most badly affected. This positive skew may again reflect measurement problems, such as changes in the testing regime. However, it could also be due to the non-normality of community transmission. For example, the number of transmissions per person is known to have a long tail, due to the presence of ‘superspreaders’. It has also been suggested that other delays, such as the infection period, may have a gamma distribution with a long tail (Shen, Taleb, and Bar-Yam (2020)).

However, if the skew is due to the asymmetry of the various statistical distributions that are involved in the complex web of transmission, why has the experience in the West been so different from that in the East? Although most European countries are affected by the *B*-strain rather then the original *A*-strain that broke out in China (Forster, Forster, Forster, and Renfrew (2020)), these strains are similar. The answer may lie in the behavior of people and their governments rather than that of the virus itself. Perhaps it is because Asian countries were better prepared following their experience with the *SARS* epidemic a decade earlier. Perhaps it is because, as Jenny (2020) argues, countries like China and Vietnam, where individual freedom is limited, were less hesitant and better able to limit the spread of the virus than elsewhere. However, Frey, Chen, and Presidente (2020) find that collectivist and democratic countries have both implemented relatively effective responses to the pandemic.

Whatever the reason for this asymmetry, it is clear that the classic model has failed us badly this time. This has been documented by several recent studies. For example, Marchant, Samia, Tanner, and Cripps (2020) show that forecasts from the Institute for Health Metrics and Evaluation (*IHME*) at the University of Washington, which underpin hospital resource planning in the US and are used in White House briefings, are usually overtaken by the data within a few days. The *IHME* model fits daily mortality data using the Gaussian bell curve (Murray (2020)).

Instead of trying to delve deeper into the data to try and find the reasons why so many countries have departed from the classic pattern, we take an empirical stance in this paper and develop a model that is based upon the statistical characteristics of the national time-series. We model the daily mortality data published by the European Centre for Disease Control (*ECDC*). We use these data rather than infections because of the acute public and policy interest focus on these data and because it is more difficult to find comprehensive and comparable data on infections.

We start by looking at the way that the tools developed by econometricians, to handle non-standard time-series representing economic growth and speculative bubbles in financial markets for example, can be used to analyze an epidemic. We then document the systematic failure of the logistic model in the US, Canada, Brazil and nine large West European countries using standard out-of-sample forecast tests. We review alternative statistical models and show that a model of the trend resembling the gamma probability density function is flexible enough to handle the positive skew now evident in many countries. Because it has greater flexibility to handle the initial stages of the epidemic, it also provides a better model than the logistic in countries like Brazil where the daily death toll is still increasing and countries like Denmark that do not have a positive skew. This model also performs well in post-sample forecast tests, suggesting that with a sample of just three or four weeks data, accurate two-, and for many countries, four-week projections can be made.

This analysis suggests that the dynamics of Covid-19 are primarily determined by the behavior of the population rather than the textbook behavior of the virus. That is because once the death toll began to soar, it then decelerated much faster than could plausibly be explained by the number of reported infections and the implied build up of immunity. It remains possible that this number is just the tip of an iceberg of infections, but as Dimdore-Miles and Miles (2020) demonstrate, it would need 250 unrecorded cases for every recorded case for immunity to explain the deceleration. It will require an antibody test survey to resolve this issue, but in the meantime it seems more plausible to think that the deceleration occurred not so much because the disease spread and built herd immunity but because news about its effects spread and made individuals and families take precautions, which were then reinforced in many countries by government lockdowns.

## 2 Modeling an epidemic using time-series econometrics

Econometricians are used to dealing with difficult economic and financial times series. Their data often violate the classical assumptions adopted in the statistical texts and thus need to be handled using special techniques. For example, macroeconomic data like *GDP* exhibit exponential growth and financial prices can exhibit speculative bubbles that are explosive. They may respond with long and variable lags to policy interventions and exogenous shocks. These data may be measured with error and subject to structural shifts as behavior or government policies change.

As Castle, A. Doornik, and Hendry (2020) argue, epidemiological data are fraught with similar problems. These econometricians have used the sophisticated linear trend fitting techniques to decompose the cumulative death counts. They split each series into trend and remainder terms, then project them forward and recombine them to produce a forecast for the following week. As they note, a significant fall in outcomes relative to extrapolations from such models can be an indication that policy interventions are having the desired effect. Epidemiologists use similar models to separate the noise from the trend in the time-series and use the trend to estimate the reproduction number R, the number of people an infected person is likely to infect.

However, deviations from a linear trend can occur for many other reasons. For example, as the death toll mounts and people begin to worry about the virus and its consequences, they are likely to modify their behavior in a way that reduces outcomes relative to a linear extrapolation. Longer term, the trend should bend as immunity builds up and the population becomes less susceptible to the disease. These endogenous feedback effects are built into the non-linear dynamics of the large-scale epidemiological models, which allow the trend to change as the epidemic progresses. This should in principle improve forecasts beyond the weekly horizon and make it more likely that systematic forecast errors are due to government interventions or other external influences.

### 2.1 The logistic process

These epidemiological models range from the large-scale computer models built by the Imperial College and other modelling groups to simple data-based curve-fitting techniques. For example, many epidemiologists fit a logistic curve to the cumulative number of infections *C*(*t*):

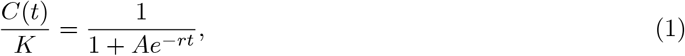

where: *A* = *K*/*C*(0) – 1, *K* is the final epidemic size and r ≥ 0 the propagation or infection rate (see for example Batista (2020), equation (2)). However, in view of the well known issues around estimation with non-stationary data Sims (1980) we model the number of new cases. Differentiating (1) and substituting Ae^−rt^ = *K*/*C*(*t*) – 1 shows that the number of new cases at any time is a bell-shaped function of the accumulated cases:

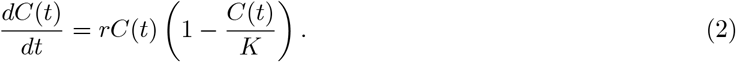

This model provides a simple way of allowing for the non-linear feedback mechanisms, loosely based on the *SIR* (susceptible, infectious, removed) model (Kermack, McKendrick, and Walker (1927), Avery, Bossert, Clark, Ellison, and Ellison (2020), Dimdore-Miles and Miles (2020)). Initially, with *C*(0) cases observed when the outbreak is detected, all of them are ‘infectious’ and will infect other ‘susceptible’ people at the rate r per unit of time (*dt*) causing *dC*(0) = *rC*(0)*dt* new cases. Thus initially, the disease will spread exponentially, at the reproduction rate *ρ* = *dC*(*t*)/(*C*(*t*)*dt*) = *r*. However, various negative feedbacks then arise, which reduce the reproduction rate.

The classic feedback mechanism is provided by herd immunity. If people who have had the disease are less susceptible to catching it again, then they move into the ‘removed’ class. As they increase as a share of the population (*N*) the probability that an infectious person will meet a susceptible one falls from 1 to (1 – *C*(*t*)/*N*)). This results in *rC*(*t*)(1 – *C*(*t*)/*N*)*dt* new cases per unit of time, resulting in (1) with *K* = *N*. However, there is a problem with this interpretation. If this were the only mechanism at work, we would expect *K* to be of a similar size to the population *N*. But it is much smaller than *N* empirically, suggesting that *C* is under-recorded. For example, Dimdore-Miles and Miles (2020) assume that the number of new cases that are symptomatic and recorded is a fraction *π* of the true number. If *C* represents the true number and *C*° the recorded number, then substituting *C* = *C*°/*π* into (2) gives the model:

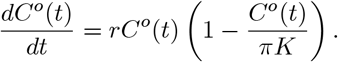

Thus the estimator *π K* effectively replaces *K*. However, as they conclude the value of *π* would need to be extremely low to fully explain this finding.

Behavioral feedbacks can also help to reduce the reproduction rate, as argued in the introduction. For example, as *C* grows, people are likely to modify their behavior in a way that mimics the effect of immunity, reducing the reproduction rate ρ = *rC*(*t*)(1 – *C*(*t*)/*K*) via the *K* parameter. This behavior can be reinforced by government interventions like lockdown. On a more pessimistic view, if immunity from exposure to the disease is partial or tends to fall with the time since exposure, then there may not be an upper limit to the cumulative number of cases.

The logistic model is designed to explain the transmission of a virus within a closed community. But apart from the country where the virus originates, all the initial cases must involve people that have recently entered the country and the number of new cases *n* will be related to the number of new passenger arrivals rather than *C*. Thus in the initial stages, before community transmission begins: *dC*(0) = n*dt* and not *dC*(0) = *nC*(0)*dt* as implied by the logistic model.

A more obvious problem with this model is that the bell and logistic curves are symmetric. The bell curve has a single peak at *C* = K/2. Once this is passed, the number of new cases begins to fall, following a path that mirrors the upward climb, before slowing to a stop as *C* approaches K. This was in fact the experience of China and many East Asian countries, which is why the logistic curve fits their data well. Unfortunately, the experience in Italy and Spain has been very different. The number of deaths fell back from the peak, but then remained stubbornly high.

### 2.2 A long, thin-tailed epidemic

We need a more flexible model to allow for these possible effects. Mathematically, we can achieve this simply by raising the *C* and (1 – C/*K*) terms in (2) by the powers *α* and *β*. This makes it a beta function, which is much more flexible:

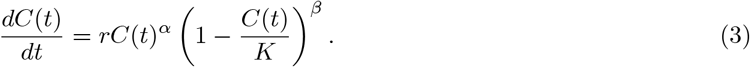

However, we find that this function is too flexible in practice, making it difficult to estimate its parameters reliably given the relatively short data sets currently available. For example, *β* usually takes a large value, producing a long but very thin tail, which makes it hard to pin down the upper bound *K*.

Instead, suppose that precautionary behavior by individuals, family groups or government means that the reproduction rate g falls exponentially as the number of cases mounts: ρ(*t*) = *re*^−γC(*t*)^. Then:

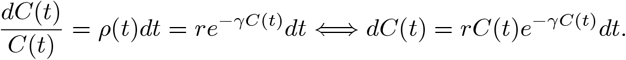

We find that although this model has two fewer parameters than the beta model, it gives a similar fit. However, its performance can be improved by changing the power of the *C* term to α, thus giving the trend a form similar to that of the gamma density function:

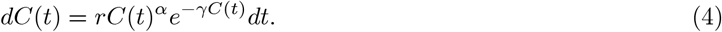

This function does not impose an upper limit on the cumulative number of cases and is still flexible enough to provide a local approximation to a bell curve and an upper skew, depending upon its parameters. It is used extensively in statistics to describe probability distributions for this reason, the well-known χ^2^ distribution being a special case (Mood, Graybill, and Boes (1973)). This function is also attractive since it can be integrated to get a closed form for the cumulative count C, the analogue of the logistic function (1) associated with the bell curve (2). Its mathematical properties are reviewed in Appendix 1.

These processes are non-stationary and should be handled using techniques developed for modelling non-stationary economic data, like growth and inflation. Their dynamics are dictated by stochastic differential equations with drift (i.e. trend) and volatility terms, like those used to model interest rates (Ait-Sahalia (1996)). We give the volatility term a form that is congruent with the drift.

## 3 Modeling the ECDC death data

The logistic model outlined in the previous section was originally developed to explain the number of new infections. However, in the absence of mass testing, the *true* numbers of people who are infected and those that have recovered are likely to be much larger than those recorded, especially if there is a large proportion of asymtomatic cases that are not recorded. To avoid these measurement problems, we extend the reasoning of the logistic infections specification to track deaths instead, following Murray (2020) and many others.

Suppose that deaths represent a constant lagged fraction of the *true* number of infections *C*(*t*). Substituting this into (4) and suitably reinterpreting the parameters, we assume:

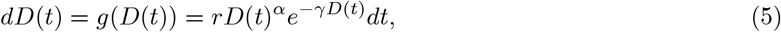

where *D*(*t*) represents the cumulative number of deaths at time *t*. To model the daily data we add a congruent volatility specification and discretize the model:

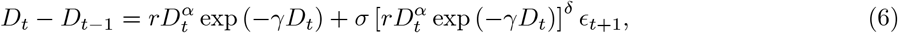

where α, γ, *r* and σ are parameters to be estimated and *ε_t_* ∼ *N* (0, 1) is a Gaussian error term. Similarly, the discrete time logistic (bell) model is

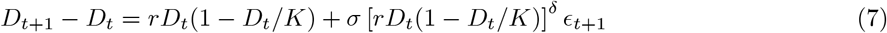

and the beta model corresponding to (3) is:

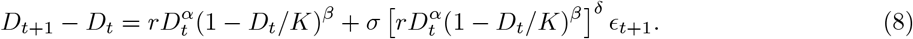

Setting α, *β* = 1 simplifies this to the logistic specification. Initial experiments showed that δ was close to 0.75 for these models for most countries and this parameter is fixed at this value in the regressions reported here.

### 3.1 China

We start by fitting three candidate models (logistic, beta and gamma) to the Chinese mortality data. Table 1 shows the regression results for the three rival models. To avoid the bias in estimates caused by an integer data count and the long left tail seen in some countries, we start the estimation for each country from the date when the cumulative number of deaths reached 15 deaths (see tables’ footnotes for more details). China was of course the first country to be hit by Covid-19 and managed to suppress it effectively by the end of March, therefore we end the sample period on 31 March 2020. Figure 2 shows the in-sample fit of the logistic (red line), gamma (black line) and beta (green line) regression models. As noted in the introduction, the bell-logistic model represents the behavior of this outbreak nicely, although the beta-drift model is better in terms of statistical criteria, and with *β* > *α*indicates a small positive skew in the data. The improvement of the fit of the beta model over the bell-logistic model is, however, achieved by construction, since since the bell model is nested by the beta model.

**Figure 2:**
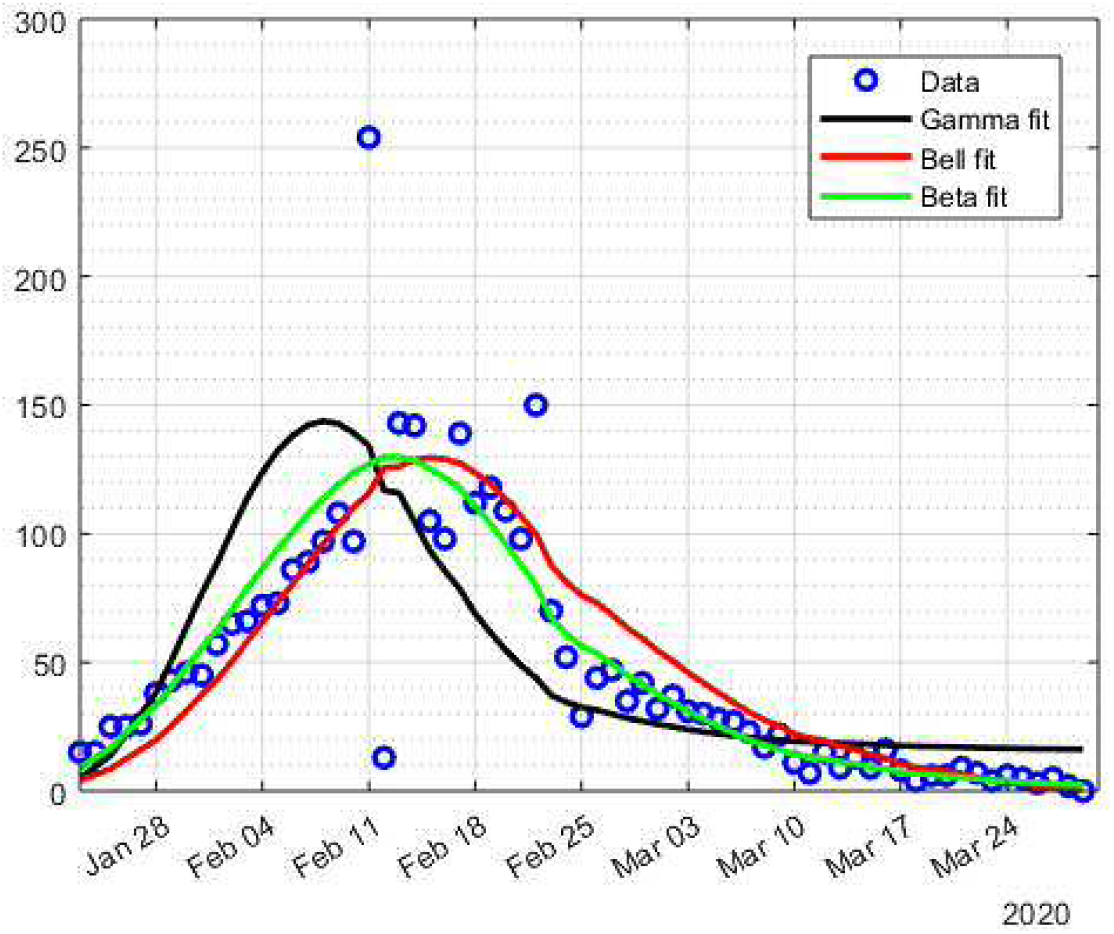
In sample fit and out of sample forecasts of daily deaths for China. The dots show data for the daily death tolls. The black line shows the regression fits for the gamma model, the red line for the logistic model and the green line for the beta model.

**Table 1:** Estimates of the logistic, gamma and beta models. Standard errors are reported in small font. The estimation period is from 23 January to 31 March 2020.

| Model | log-lik. | $r$ | $K$ | $\alpha$ | $\beta$ | $\gamma \times 10,000$ | $\sigma$ |
| --- | --- | --- | --- | --- | --- | --- | --- |
| China |  |  |  |  |  |  |  |
| Bell | -284.98 | 0.1566<br>0.0100 | 3,309<br>3 | - | - | - | 1.2981<br>0.1283 |
| Gamma | -310.45 | 0.0689<br>0.0388 | - | 1.3384<br>0.1107 | - | 16.290<br>1.494 | 1.5828<br>0.1683 |
| Beta | -257.40 | 0.5820<br>0.1747 | 3,349<br>19 | 0.8387<br>0.0505 | 1.2343<br>0.1007 | - | 0.8537<br>0.0789 |

### 3.2 Italy and Spain

We next apply these three models to Italy and Spain, which were the first western countries to be overwhelmed by the virus and where the skew in the mortality figures first became apparent. Table 2 shows the regression results for the three rival models. The poor performance of the logistic bell curve is apparent from the likelihood statistics, which show that the gamma and beta models fit the data much better. Moreover, the estimates of *K*, which indicates the final size of the epidemic, look very low, only slightly ahead of the cumulative number of deaths as they stand at the time of writing. This is because the bell curve predicts a sharp deceleration, mirroring the initial explosive acceleration, which has regrettably proved to be too optimistic. The beta model predicts a much larger final death toll in Italy. However, this estimate is poorly defined, since as noted, the large value of the power parameter *β* means the tail is very thin, making it hard to know where to truncate it. This model is clearly over-parameterised. The gamma model is much more robust. It has one fewer parameters and these parameters are estimated more precisely.

**Table 2:**
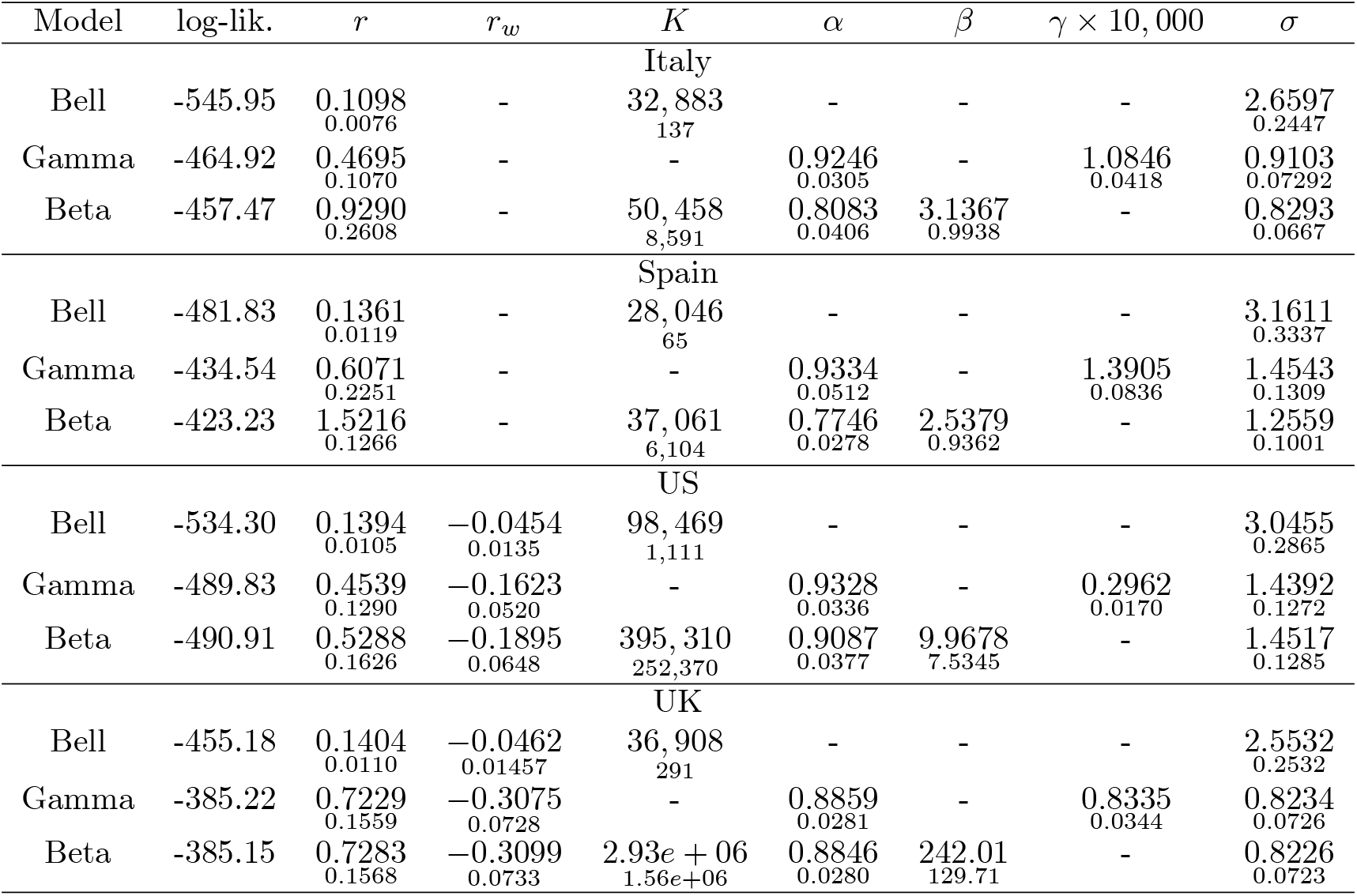
Estimates of the logistic, gamma and beta models. Standard errors are reported in small font. The estimation periods start on: Italy – 29 February, Spain – 11 March, US – 9 March, UK – 16 March and ends on 20 May 2020, except for Spain – 19 May 2020.

The in-sample fit of the logistic and gamma regression models for Italy and Spain is shown in the panels on the left hand side of Figure 3. These figures make it clear that the logistic function performs poorly. Its reproduction rate is initially too slow, which means that the predicted peak comes too late and is followed by a decline in mortality that is too rapid. The gamma model, on the other hand, plots straight through the observations. The dashed lines in the left panels of Figure 3 show the median out-of-sample forecast for both models based on simulations for the period of four weeks following the last observation. The shaded areas indicate the confidence intervals for the gamma model. The logistic model predicts that the daily death rates for both countries will decline to close to zero within a couple of weeks, while the gamma model indicates that the number of deaths will continue to hover around 150 – 200, declining slowly. Appendix 2 explains the simulation procedure.

**Figure 3:**
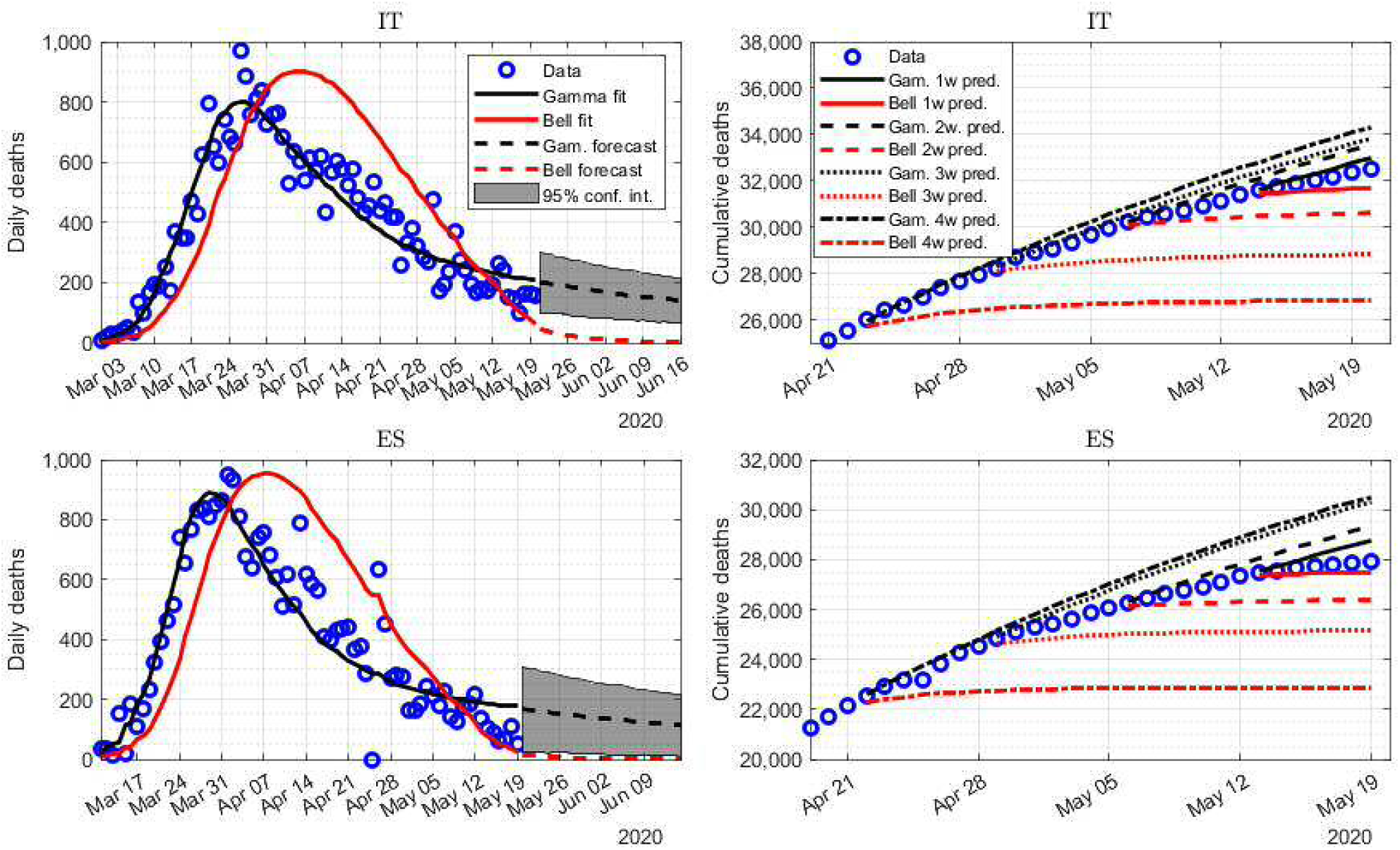
In sample fit and out of sample forecasts of daily deaths for Italy and Spain. The dots show data for the daily (left panels) and cumulative (right panels) death tolls. In the left panels, the continuous lines show the regression fits for the logistic (red) and gamma (black) models. The dashed lines show the forecast made for the next four weeks with the gamma-drift model using the full sample estimates conditional upon the last observation in the sample, together with the confidence intervals. The right hand panels show out-of-sample forecasts based on the parameters estimated on samples ending one (continuous lines), two (dashed lines), three (dotted lines) and four (dash-dotted lines) weeks before the most recent observation.

The tendency for the logistic bell curve model to under-predict the final death toll is brought out more clearly by right-hand side panels of Figure 3. These illustrate out-of-sample dynamic forecasts based on the parameters estimated on samples ending one (continuous lines), two (dashed lines), three (dotted lines) and four (dash-dotted lines) weeks before the most recent observation 20 May 2020.^3^ For instance, the continuous lines show the forecasts of the logistic (red) and gamma (black) models estimated on 13 May for the three week period ending on 20 May. It shows that the logistic model tends to under-predict the most recent observations. It invariably predicts a sharp fall in the death count that is almost immediately overtaken by the out-turn. This is also the case with the two- three- and four-week ahead forecasts. Remarkably, despite these relatively short data sets, the gamma model predicts the behavior of the mortality data in these two countries nicely over this period, predicting a gradual fall in the daily counts and the slope of the cumulative curve. In strong contrast, the logistic bell model tends to flatten out immediately on, or shortly after the date the forecast was made. The beta model is not used in this exercise because its parameters are often poorly determined in short data samples, which can cause it to forecast erratically out of sample.

### 3.3 The US and the UK

Next we turn to the US and the UK, two of the countries with the highest total number of deaths. The daily counts in these countries are also proving to be stubbornly high. Due to operational issues in these countries, the reported number of Covid-19-related deaths is significantly lower on weekends than on weekdays. We account for this feature of data by interacting the rate parameter r with a dummy variable for a weekend day, so for instance, we estimate the gamma model as:

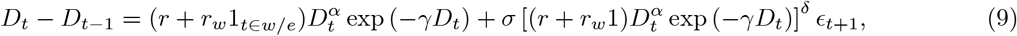

where 1_*tεm/e*_ is an indicator variable equal to 1 if the day *t* is a weekend day, and zero otherwise. We also incorporate the weekend effect in the logistic and gamma models for both countries.

The regression results for the US and the UK are shown in Table 2. The poor performance of the bell curve is again apparent from the likelihood statistics and the low estimates of *K*. The beta model is clearly over-parameterised and gives a nonsensical estimate of *K*. Nevertheless, the gamma model has a likelihood similar to that of the beta model and its parameters are nicely identified.

The left-hand side panels of Figure 4 shows the fit of the logistic and gamma models to the daily death rates for the US and the UK. Again, the peak of the epidemic is again too late in the logistic model, although the misalignment with the observations is not as pronounced as it is for Italy and Spain. Although the weekend dummy brings the logistic model closer to the weekend data, these predictions are far from close. The gamma model, on the other hand, closely fits both the weekday and weekend data. The dashed lines show the out-of-sample forecasts for the next three weeks. The logistic model predicts a rapid decline in death rates to negligible levels over this period, while the gamma model forecasts death rates of 500 – 1, 000 for the US and 200 – 400 for the UK.

**Figure 4:**
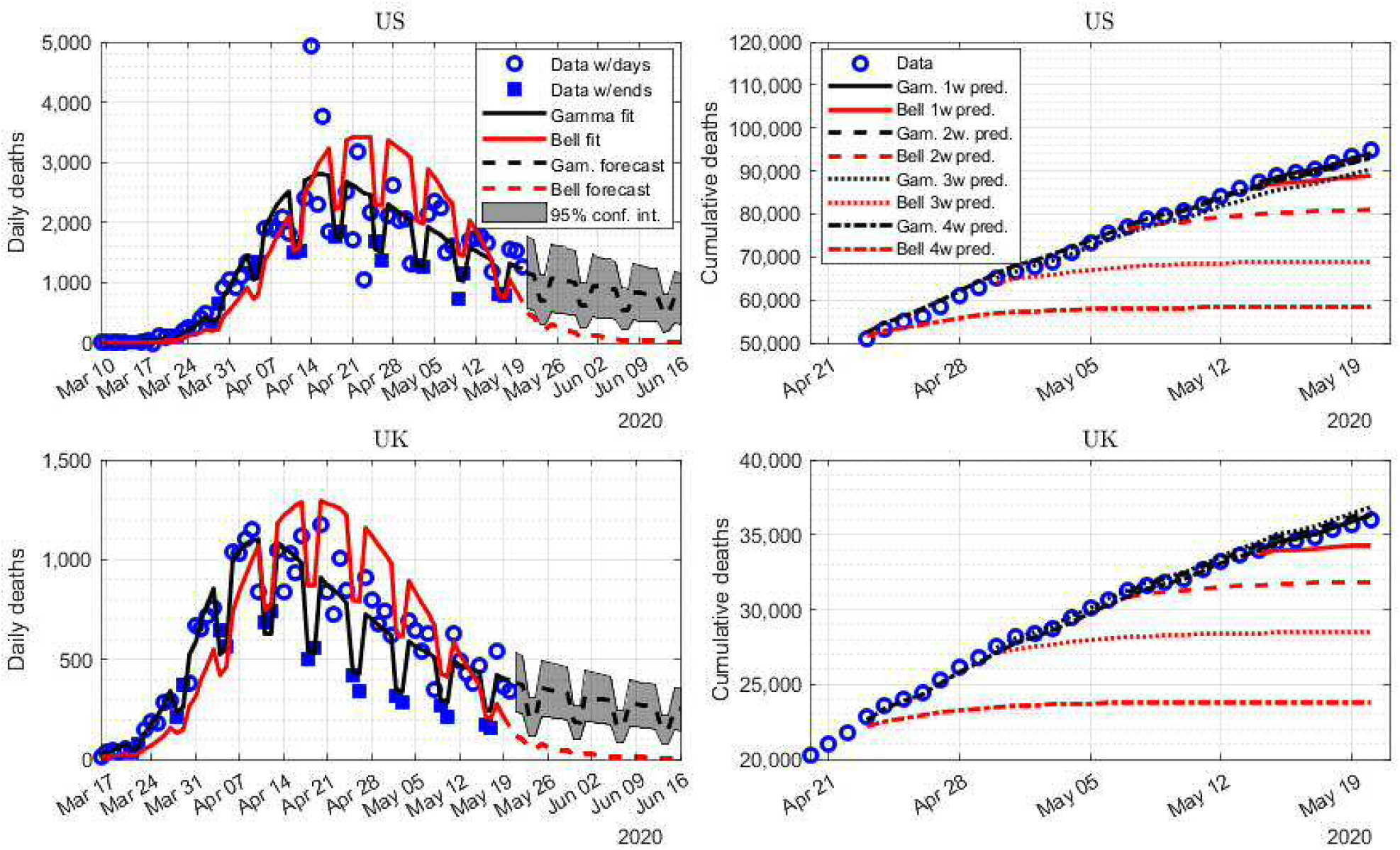
In sample fit and out of sample forecasts of daily deaths for the US and the UK. In the left hand panels, the dots show weekday observations of the daily death toll and the squares show observations that are affected by the weekend effect. The continuous lines show the regression fits for the logistic (red) and gamma (black) models. The dashed lines show the forecast made for the next four weeks with the gamma-drift model using the full sample estimates conditional upon the last observation in the sample, together with the confidence intervals. In the right hand panels, the dots show data for the cumulative death toll. Out-of-sample forecasts are shown for samples ending one (continuous lines), two (dashed lines), three (dotted lines) and four (dash-dotted) weeks before the most recent observation.

The right-hand panels of Figure 4 again shows the persistent tendency of the logistic curve to under-predict the total number of deaths. On the other hand, the out-of-sample-sample forecasts of the gamma model for the US are closely aligned with the data out-turn up to a month ahead, as are the forecasts for the UK.

One feature that stands out from this table is that for both the beta and (with the exception of China, Belgium and France) gamma models, the estimates of *α*are consistently below unity. This feature helps these models to handle the initial effects of the virus, compared with the logistic which imposes *α*= 1. This is because estimates of *α*are largely determined by initial stages of the epidemic, while *β* and γ are more strongly influenced by the post-peak behavior. As noted in Section 2.1, the first stage will be driven by infected people entering the country at the rate n*dt* rather than reproduction in the community. Strictly speaking, the community transmission statistic ρ = *dC*/*C* = *ndt*/*nt* = *dt*/t is then meaningless, but runs to a larger and larger number as we go back in time. This is consistent with the behavior of the gamma model with 0 < *α*< 1, which we can approximate by ρ = *dC*/*C* = *C*^*α*— 1^*dt* close to *C* = 0.

### 3.4 Other countries

One of the notable features of the current crisis is that the six Eastern European accession states have remained largely untouched by the virus. This is also true of countries like Portugal, Greece, Slovenia and Croatia, that joined the EU after the initial expansion in 1973, though not of course Spain. Nevertheless, most Western European countries have been affected in varying degrees. We estimate the models for seven other West European countries affected by Covid-19 to a varying degree: Belgium, Denmark, France, Germany, Ireland, the Netherlands and Sweden. We also estimate models for Canada and Brazil. We found significant weekend effects in the Netherlands and Sweden. The regression results for these two countries are reported in Table 4 and for the other countries in Table 3.

**Table 3:**
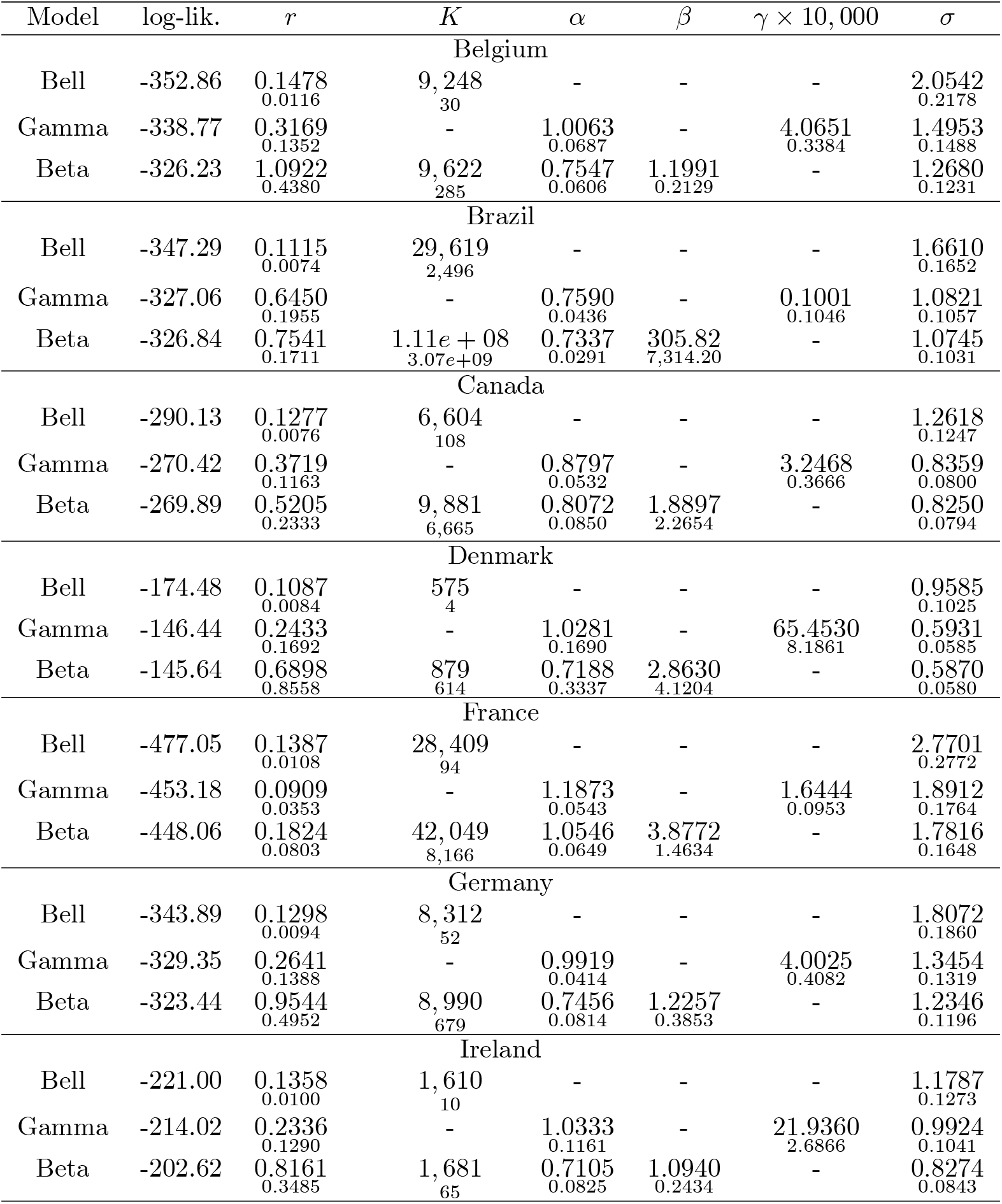
Estimates of the logistic, gamma and beta models. Standard errors are reported in small font. The estimation periods start on: Belgium – 31 March, Brazil – 23 March, Canada – 23 March, Denmark – 25 March, France – 10 March, Germany – 21 March, Ireland – 28 March 2020 and ends on 20 May 2020.

| Model | log-lik. | $r$ | $K$ | $\alpha$ | $\beta$ | $\gamma \times 10,000$ | $\sigma$ |
| --- | --- | --- | --- | --- | --- | --- | --- |
| Belgium |  |  |  |  |  |  |  |
| Bell | -352.86 | 0.1478<br>0.0116 | 9,248<br>30 | - | - | - | 2.0542<br>0.2178 |
| Gamma | -338.77 | 0.3169<br>0.1352 | - | 1.0063<br>0.0687 | - | 4.0651<br>0.3384 | 1.4953<br>0.1488 |
| Beta | -326.23 | 1.0922<br>0.4380 | 9,622<br>285 | 0.7547<br>0.0606 | 1.1991<br>0.2129 | - | 1.2680<br>0.1231 |
| Brazil |  |  |  |  |  |  |  |
| Bell | -347.29 | 0.1115<br>0.0074 | 29,619<br>2,496 | - | - | - | 1.6610<br>0.1652 |
| Gamma | -327.06 | 0.6450<br>0.1955 | - | 0.7590<br>0.0436 | - | 0.1001<br>0.1046 | 1.0821<br>0.1057 |
| Beta | -326.84 | 0.7541<br>0.1711 | 1.11e+08<br>3.07e+09 | 0.7337<br>0.0291 | 305.82<br>7,314.20 | - | 1.0745<br>0.1031 |
| Canada |  |  |  |  |  |  |  |
| Bell | -290.13 | 0.1277<br>0.0076 | 6,604<br>108 | - | - | - | 1.2618<br>0.1247 |
| Gamma | -270.42 | 0.3719<br>0.1163 | - | 0.8797<br>0.0532 | - | 3.2468<br>0.3666 | 0.8359<br>0.0800 |
| Beta | -269.89 | 0.5205<br>0.2333 | 9,881<br>6,665 | 0.8072<br>0.0850 | 1.8897<br>2.2654 | - | 0.8250<br>0.0794 |
| Denmark |  |  |  |  |  |  |  |
| Bell | -174.48 | 0.1087<br>0.0084 | 575<br>4 | - | - | - | 0.9585<br>0.1025 |
| Gamma | -146.44 | 0.2433<br>0.1692 | - | 1.0281<br>0.1690 | - | 65.4530<br>8.1861 | 0.5931<br>0.0585 |
| Beta | -145.64 | 0.6898<br>0.8558 | 879<br>614 | 0.7188<br>0.3337 | 2.8630<br>4.1204 | - | 0.5870<br>0.0580 |
| France |  |  |  |  |  |  |  |
| Bell | -477.05 | 0.1387<br>0.0108 | 28,409<br>94 | - | - | - | 2.7701<br>0.2772 |
| Gamma | -453.18 | 0.0909<br>0.0353 | - | 1.1873<br>0.0543 | - | 1.6444<br>0.0953 | 1.8912<br>0.1764 |
| Beta | -448.06 | 0.1824<br>0.0803 | 42,049<br>8,166 | 1.0546<br>0.0649 | 3.8772<br>1.4634 | - | 1.7816<br>0.1648 |
| Germany |  |  |  |  |  |  |  |
| Bell | -343.89 | 0.1298<br>0.0094 | 8,312<br>52 | - | - | - | 1.8072<br>0.1860 |
| Gamma | -329.35 | 0.2641<br>0.1388 | - | 0.9919<br>0.0414 | - | 4.0025<br>0.4082 | 1.3454<br>0.1319 |
| Beta | -323.44 | 0.9544<br>0.4952 | 8,990<br>679 | 0.7456<br>0.0814 | 1.2257<br>0.3853 | - | 1.2346<br>0.1196 |
| Ireland |  |  |  |  |  |  |  |
| Bell | -221.00 | 0.1358<br>0.0100 | 1,610<br>10 | - | - | - | 1.1787<br>0.1273 |
| Gamma | -214.02 | 0.2336<br>0.1290 | - | 1.0333<br>0.1161 | - | 21.9360<br>2.6866 | 0.9924<br>0.1041 |
| Beta | -202.62 | 0.8161<br>0.3485 | 1,681<br>65 | 0.7105<br>0.0825 | 1.0940<br>0.2434 | - | 0.8274<br>0.0843 |

**Table 4:** Estimates of the logistic, gamma and beta models. Standard errors are reported in small font. The estimation periods start on: the Netherlands – 17 March, Sweden – 22 March and ends on 20 May 2020.

| Model | log-lik. | $r$ | $r_w$ | $K$ | $\alpha$ | $\beta$ | $\gamma \times 10,000$ | $\sigma$ |
| --- | --- | --- | --- | --- | --- | --- | --- | --- |
| The Netherlands |  |  |  |  |  |  |  |  |
| Bell | -339.58 | 0.1440<br>0.0116 | -0.0522<br>0.01512 | 5,826<br>22 | - | - | - | 1.7217<br>0.1747 |
| Gamma | -301.89 | 0.5178<br>0.1853 | -0.2090<br>0.0798 | - | 0.9269<br>0.0606 | - | 6.0142<br>0.4201 | 0.9059<br>0.0830 |
| Beta | -293.24 | 1.1433<br>0.2226 | -0.4473<br>0.1100 | 7,623<br>1,090 | 0.7557<br>0.0118 | 2.2104<br>0.7142 | - | 0.8045<br>0.0686 |
| Sweden |  |  |  |  |  |  |  |  |
| Bell | -305.35 | 0.1408<br>0.0148 | -0.0678<br>0.0167 | 4,012<br>61 | - | - | - | 1.9371<br>0.2181 |
| Gamma | -292.27 | 0.4383<br>0.2587 | -0.2262<br>0.1405 | - | 0.8911<br>0.1064 | - | 6.4975<br>1.0913 | 1.4761<br>0.1557 |
| Beta | -292.33 | 0.4937<br>0.1949 | -0.2549<br>0.1119 | 19,332<br>56,593 | 0.8630<br>0.0375 | 10.896<br>36.856 | - | 1.4763<br>0.1555 |

The beta model performs as well or better than the gamma model for these countries in terms of likelihood. Moreover, the parameter *β* is generally not significantly different from unity, showing that the marked outperformance against the logistic model is due to the relaxation of the unit restriction on and *α*not *β*. Nevertheless, the gamma model is much better determined in the shorter data sets used for the post-sample forecast tests and we focus on that model in what follows. Figures 6, 5 and 7 show the results of the post-sample tests. Once again, the tendency for the logistic model to systematically under-predict the recent data is evident. The gamma model generally provides accurate out-of-sample forecasts, as far as four weeks ahead for many countries. One exception here is Ireland, where the four-week ahead prediction from the gamma model tends to overshoot.^4^ The parameters of the gamma model are poorly determined on this short sample and so this result should be ignored.

**Figure 5:**
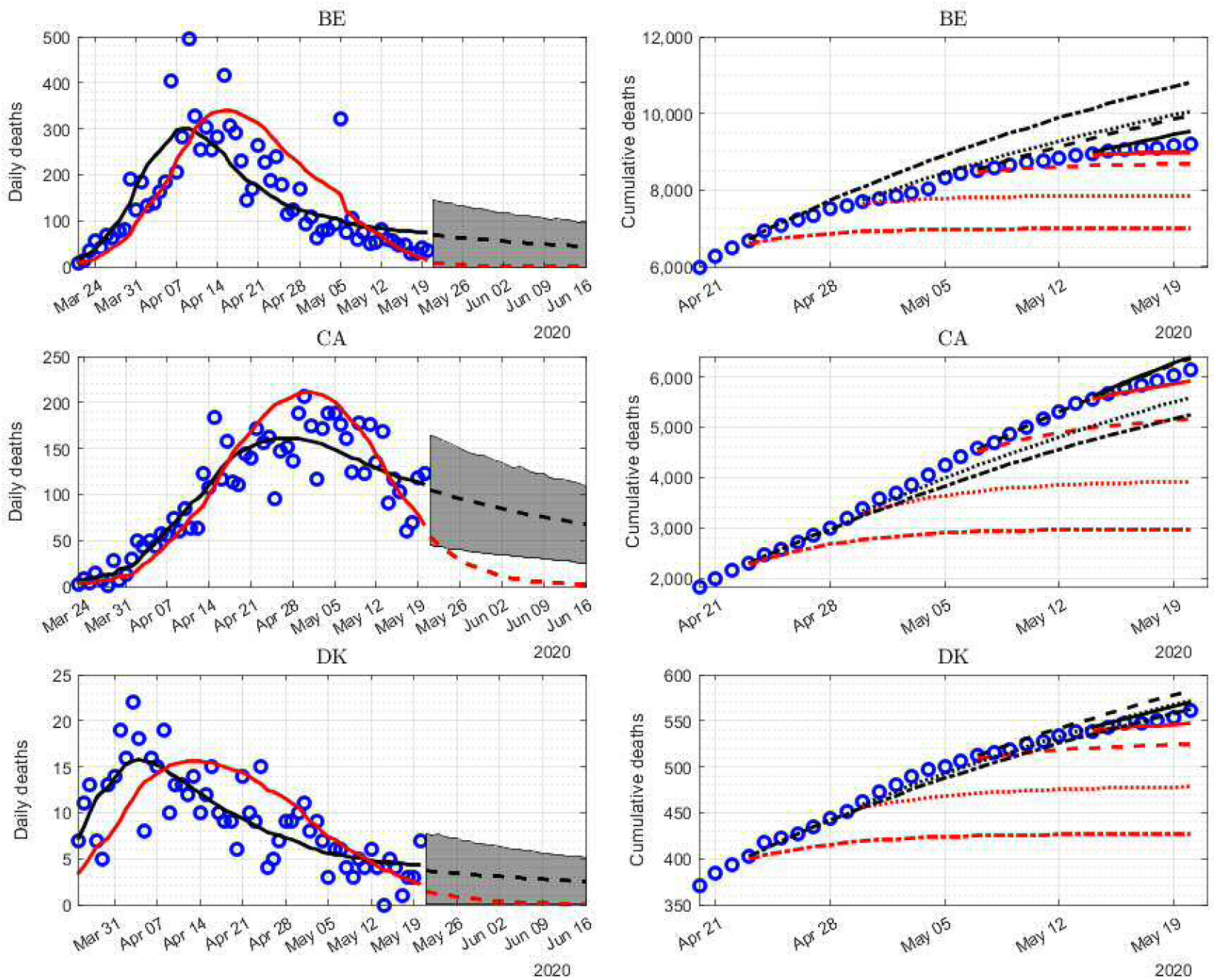
In sample fit and out of sample forecasts of daily deaths for Belgium, Canada and Denmark. See notes toFigure 3.

**Figure 7:**
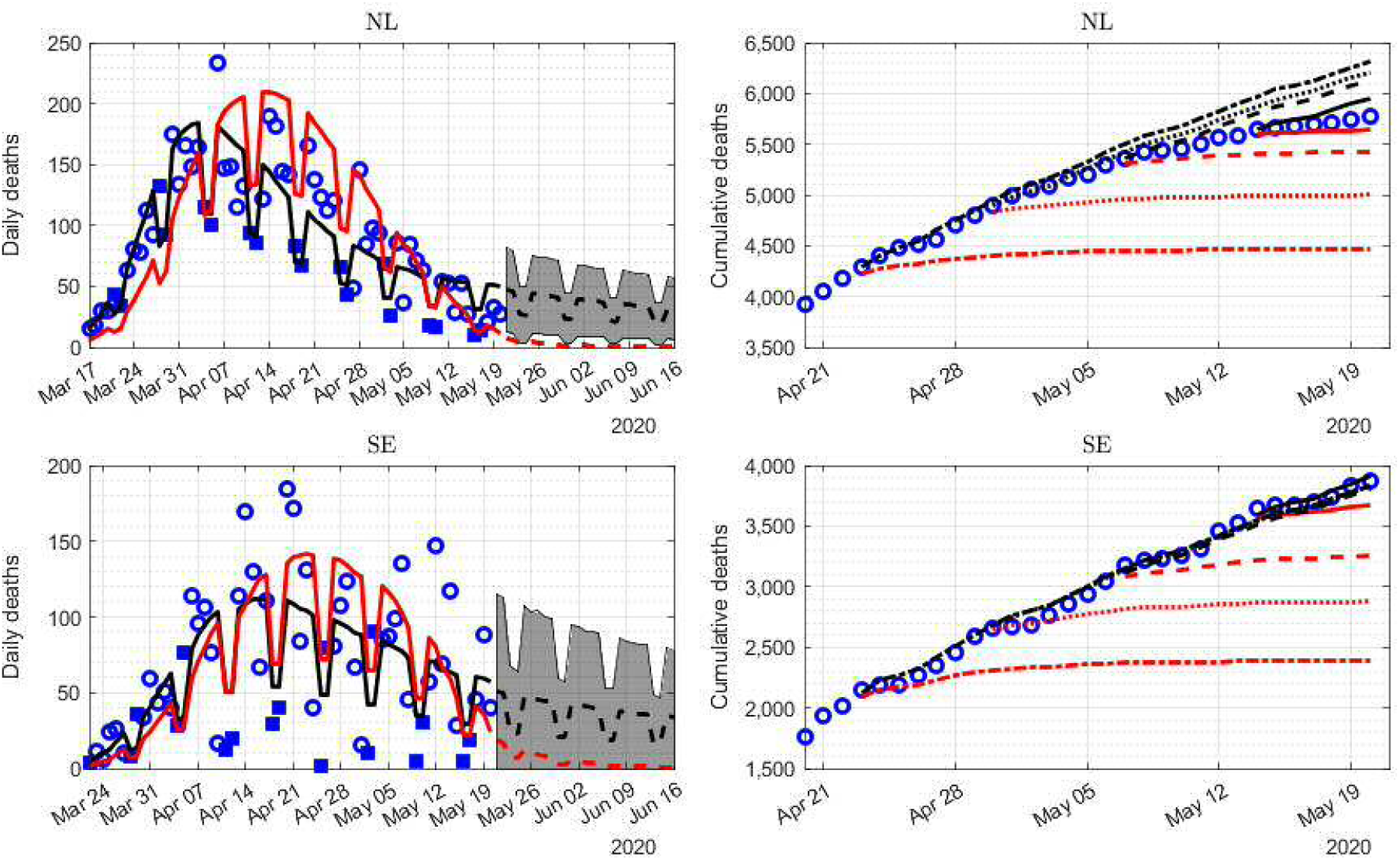
In sample fit and out of sample forecasts of daily deaths for the Netherlands and Sweden. See notes toFigure 4.

Germany, Denmark and Sweden provide interesting special cases. Germany has successfully combined a lockdown with mass population testing. Denmark was the second European country after Italy to go into lockdown, on 11 March, before any fatalities had occurred. Sweden is exceptional in having relied on individual responsibility rather than lockdown to contain the spread of the virus. Nevertheless, the mortality rate (deaths relative to population) is no higher than the median. Born, Dietrich, and Gernot Müller (2010) use synthetic control techniques to construct a doppelganger for Sweden on the basis of pre-lockdown infection rates and show that the Swedish dynamics are no difference post-lockdown. Our time-series work shows that although the bell curve model provides a satisfactory fit for Sweden, it systematically under-forecasts, just as it does for other Western countries.

Another interesting case is Brazil, which is still experiencing a rising daily death toll. Despite this, the parameters of the gamma model for Brazil are precisely estimated. The forecast formulated four weeks before the end of the sample (see Figure 8) under-predicts in the long run, but still provides much accurate prediction than the bell model, especially in the one or two weeks horizon. The other within-sample forecasts, one-, two- and three-weeks ahead predict the dead toll very accurately. The bell model, in contrast, consistently predicts that the end of (sub)sample is close to the epidemic peak and the total death number that is overtaken by the out-turn within a couple of days.

**Figure 8:**
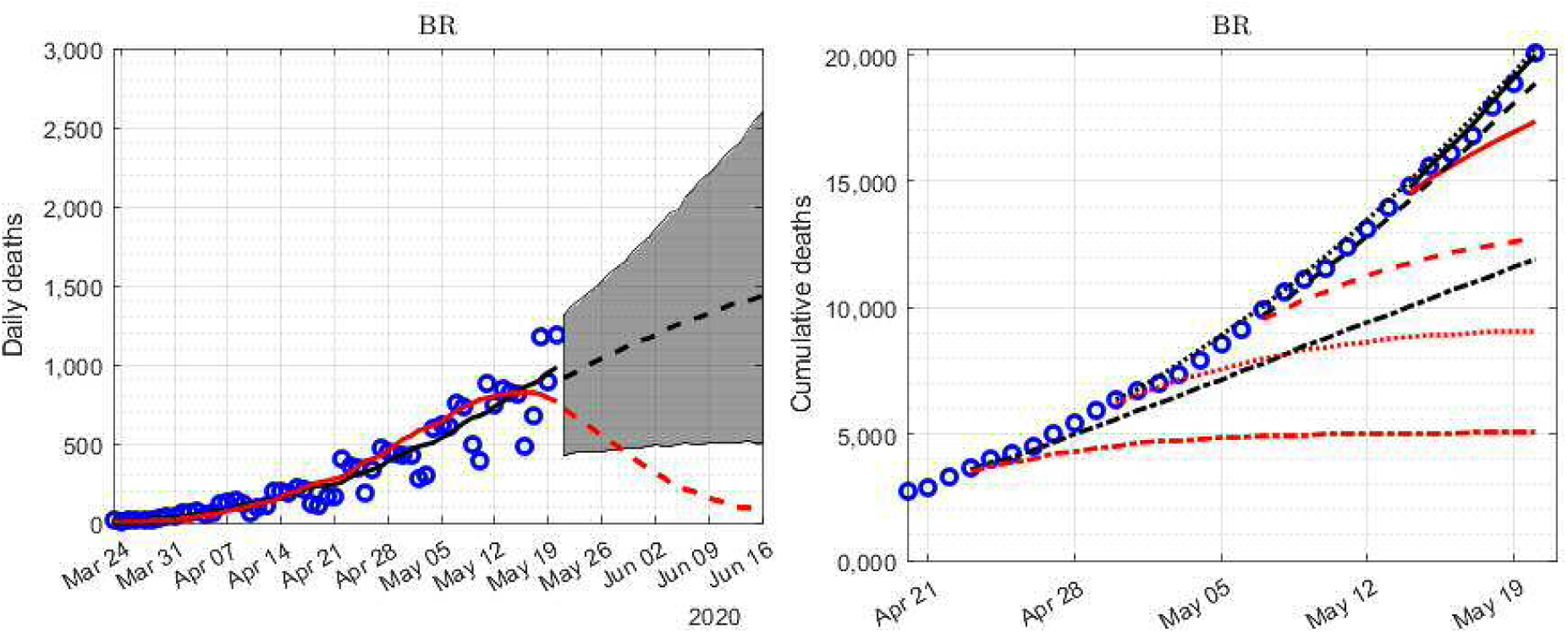
In sample fit and out of sample forecasts of daily deaths for Brazil. See notes toFigure 3.

### 3.5 Statistical comparisons

Time series models are designed to abstract from the noise in the data and provide estimates of the trend in the series. In the case of non-linear process like an epidemic, they can also be used to indicate the rate at which the trend is increasing or decreasing and whether this rate is accelerating or decelerating. In this case, the trend in the cumulative death toll is the estimated number of daily deaths, described by the gamma drift function in equations (8) or (9). Its properties can be seen from the shape of the curves in Figures 3 to 8. However, rather than ‘eyeballing’ charts it is often better to look at the results numerically, using well-known statistics, particularly when comparing different countries. Table 5 shows some of the basic numbers that emerge from this study for the US and Canada and West European countries. These arguably allow a broader assessment of the characteristics of the virus than comparisons of death tolls in different countries.

**Table 5:**
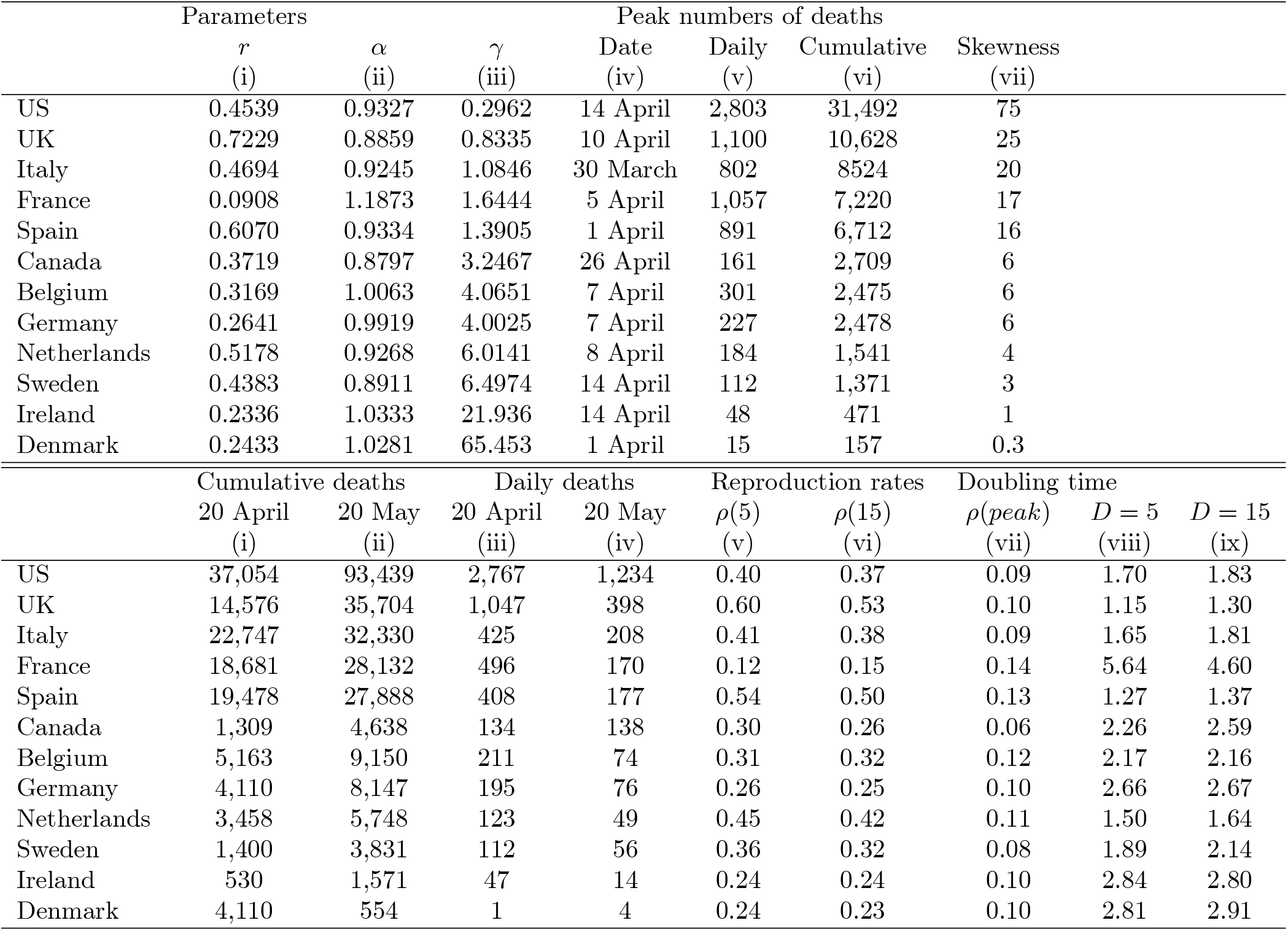
Summary statistics for gamma drift models.

These statistics follow from the well-known mathematical properties of gamma-type functions, which are reviewed in Appendix 1. The first three columns show the parameters estimated for each country, reproduced from Tables 2 to 4. These are first used to determine when the peak in the death toll is likely to have occurred. The table shows the date that this was reached in each country; the estimated number of daily deaths at the peak (corresponding to the height of the peak in each figure) and the cumulative number of deaths at that point.

These parameters are then used to calculate the ‘skewness’ coefficient for each country, shown in column (vii). This indicates how different the decline from the peak is proving to be, compared to the rise from the first few cases to the peak. The rows of this table are ranked in terms of skewness, starting with the US at the top. These statistics indicate a clear divide between the US, the UK, Italy, Spain and France on the one hand with a marked positive skew and other West European countries on the other, with hardly any skew. Clearly, the outperformance of gamma and beta models for this second group is not due to a long upper tail, but due to its ability to handle the start of the epidemic and other characteristics. The second panel of the table gives the latest numerical estimates from the model and compares these with the values implied by these full sample parameters a month earlier. These calculations depend upon the cumulative number of deaths at these times, shown in the first two columns. Columns (iii) to (iv) show how the estimated daily death toll has fallen back since the peak in each country (shown in column (v) of the top panel).

Although this research project was initially aimed at modeling the features of the stubborn upper tail seen in Italy and elsewhere, these results also help us to a better understanding of the early, exponential, phase of this pandemic. In this early, pre-peak phase, attention is focussed on the time that it takes for the cumulative number of infections to double. One of the striking features of Tables 1 to 4 is that the estimates of the parameter ρ(1) = *r* from the bell-logistic model are remarkably close, ranging from 0.11 for Denmark to 0.157 for China. This parameter is important because it shows the daily growth rate during the initial phase of the epidemic, before the various negative feedback effects are significant. Thus the logistic model would suggest that country-specific factors like age structure, population density, social behavior and size of capital city that are widely regarded as influencing the spread of the disease are probably much less important than supposed initially. However, the gamma drift model, which allows more flexibility in the initial phase through the parameter α, suggests that the logistic model covers up important idiosyncratic effects. Table 5 shows the values of ρ(5) and ρ(15) and the respective doubling times during the initial phase. With *D* = 5 the doubling time ranges widely, from just 1.3 days for Spain to 5.6 for France, in strong contrast to the impression given by the logistic model.

## 4 Conclusion

There are many ways of analyzing the economy and the same is true of an epidemic. On the one hand, we need detailed structural models to analyze the likely effects of public health interventions like the current lock-down and on the other we need reduced-form data-based models to track the progression of the virus and assess its likely evolution. Our approach to this problem is based on non-linear stochastic difference equations that are designed to mimic the theoretical dynamics of an epidemic. Our experience reminds us that conventional models often use inappropriate parameter restrictions and that post-sample forecast performance is crucial in testing these. We find that a model that uses a gamma-type function to model the drift and volatility in the daily mortality data emerges from these tests and is tracking the evolution of this epidemic nicely, even in countries that are not following the classic pattern seen in China and many East Asian countries.

Time series models provide useful statistics that summarize the virulence, morbidity and mortality rates in different countries. We can use these to look at the effects on these indicators of variations in containment and testing strategies across a cross-section of countries, while controlling for different demographic and other characteristics. Their dynamics are the result of a convolution of the long and possibly variable lags involved in the data generation process, as they are in any model. In this case, they involve infection, reporting and other lags as well as behavioral responses that can change over time, causing systematic deviations from the projected path. These provide an early warning signal that the dynamics are changing, either because policies or people are changing or because the virus itself has changed.

Looking forward, we can potentially use these models to identify structural breaks and analyze the impact of policy interventions. Econometricians have a variety of handy tools for conducting this kind of task, including tests for discrete changes when the break-point is unknown *a priori* as well as tests for breaks at points when a change is likely to have occurred, due to policy a interventions for example. The explosive behavior test proposed in a series of papers by Phillips, Shi and Yu (Phillips, S. Shi, and Yu (2014), Phillips, S. Shi, and Yu (2015)) also looks potentially useful in this respect. This was developed for testing bubbles in financial time-series, which are analogous to the early phase of the epidemic spread.

Our daily short-term forecasts of the coronavirus mortality rates for the UK are available on our website: https://sites.google.com/york.ac.uk/adam-golinski/coronametrics. The Matlab code is available upon request.

## Data Availability

We model the daily mortality data published by the European Centre for Disease Control (ECDC).

## Appendix 1 Properties of the gamma drift function

The mathematical properties of gamma-type functions are well known. This function is used to define the gamma probability distribution Mood, Graybill, and Boes (1973). Health economists are also used to dealing with skewness and kurtosis. For example Jones, Lomas, and Rice (2015) use generalized gamma and beta functions to handle the tails in health cost distributions.

### The peak in mortality

The value of the drift in (6) at any time gives the expected number of deaths and is obtained by substituting the cumulative number of deaths at that time. However, analytically, it is often easier to work in continuous rather than discrete time, using (5) instead of (6). Gamma-type functions have a single peak. This is found by taking the first derivative of the drift:

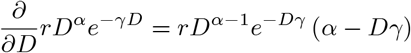

and setting this to zero by setting *D* = α/γ. These values are shown in column of the first panel of Table 5 and substituting them back into (5) gives the estimate of the daily death toll at the peak. The peak corresponds to the mode in the gamma distribution. Similarly, the skewness coefficient shown in column(vii) follows from that of the gamma distribution (Mood, Graybill, and Boes (1973)):

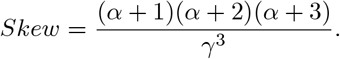

### Doubling time

Although this paper is focussed on the features of the stubborn upper tail, our results also help us to a better understanding of the early, exponential, phase of this pandemic. In that phase, attention is focussed on the time that it takes for the cumulative number of infections to double. This can also be used to assess the initial behavior of the mortality rate. But to gauge that we need to have an estimate of the reproduction rate ρ(*D*(*t*)). Table 5 shows three estimates implied by the gamma drift function (8). These show the expected daily change as a decimal fraction of the cumulative. The first is evaluated at the start of the sample with *D* = 15, suggesting that is in the range of 0.2 – 0.4 for most countries. The second is a pre-sample extrapolation value for *D* = 5. The third is the value at the peak.

We can then calculate the doubling time by dividing (5) by *D* to get the percentage or logarithmic change:

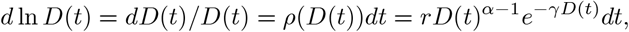

which we can integrate to get the approximation: ln *D*(*t* +τ)/*D*(*t*) = ρ(*D*(*t*))τ, where τ is measured in days.

The doubling time is found by setting *D*(*t* + τ)/*D*(*t*) = 2 and solving for τ:

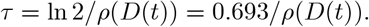

As noted in Section 3.5, the values in the table range widely across different countries. Absent a second wave, the cumulative cannot re-double once the peak is passed, at least in the case of a symmetric specification.

## Appendix 2 Out-of-sample forecast simulation

We analyze the uncertainty surrounding the out-of-sample forecasts using Monte Carlo simulations. There are two sources of uncertainty to evaluate. The first source of uncertainty concerns the parameters. This is indicated by their estimated variance-covariance matrix. To capture the correlation between the parameter estimates, we decompose the variance-covariance matrix of the estimates using the Cholesky decomposition and generate a new set of parameters as

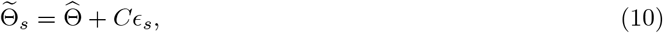

where 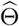 is a vector of maximum likelihood parameter estimates, *C* is the lower-triangular of the variance matrix of 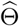 and *∊_s_* is a vector of standard normal variable generated by a random number generator.

The second source of uncertainty comes from the additive equation error term *e_t_*, geared up by the volatility term. This uncertainty builds up initially with the horizon of the forecast, but eventually starts to decline as the process exceeds the peak. Given the (randomized) parameter values, a random number generator of a standard normal variable is used to simulate values of *e_t_*, which are then fed back into the dynamics equation (6). For example, given the parameters generated for the s’th simulation 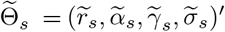, we simulate the future realizations of the gamma process for *t* = 1,…, 21 as:

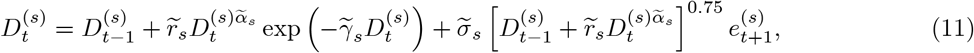

where *t* = 0 is the last observation in the sample. We repeat this procedure 10, 000 times. The daily death figures show the median and the 95% interval of these simulations.

## Footnotes

1 These data are available at: https://covidtracking.com/data.

2 These data are available at: http://www.england.nhs.uk/statistics/statistical-work-areas/covid-19-daily-deaths/. A correction for the most recent reporting delay is available at: http://users.ox.ac.uk/~nuff0078/Covid/index.htm

3 The dynamic forecast from any date is made using the parameter estimates from the sample ending on that data. The last cumulative count in this sample is used to predict the daily count for the following day, which is then added to the cumulative used to predict the next day’s count, with this procedure repeated until the forecast horizon. This provides a forecast conditional upon the sample.

4 The Irish Department of Health changed the reporting methodology on 24 April, adjusting the death toll upwards by 18τ additional cases. We account for this adjustment by re-scaling the daily death rates before the adjustement. We are grateful to Donal Smith for help with this.

